# Predicting gait patterns from actionable impairments in Duchenne muscular dystrophy: A Machine Learning and Explainable Artificial Intelligence study

**DOI:** 10.64898/2026.08.24.26361175

**Authors:** Ines Vandekerckhove, Briek Lismont, Tinne De Laet

## Abstract

**Background:** Prolonging ambulation is an important treatment goal in children with Duchenne muscular dystrophy (DMD). Clinical management targets ‘actionable’ (i.e., modifiable) impairments, such as progressive muscle weakness and contractures, that underlie gait pathology. Gait classification may improve clinical decision-making, but the utility of gait classification in clinical practice depends on understanding how underlying, actionable impairments contribute to distinct gait patterns, which remains insufficiently understood. The research questions were: (1) Can DMD gait patterns be accurately classified from actionable impairments? and (2) Can the model’s predictions be explained, and do these explanations provide clinical utility and increase trust in the model?

**Methods:** A retrospective dataset of 274 lower-limb observations from 137 assessments in 30 boys with DMD was analyzed, including 3D gait analysis, instrumented strength assessment, and clinical examination (manual muscle testing, goniometry and clinical stiffness scale). Observations were classified into the mildly affected, tiptoeing, or flexion gait pattern. Ten predictors representing actionable impairments were included: nine predictors related to muscle weakness and contractures, and body mass index (BMI). A balanced random forest classifier was evaluated with leave-one-group-out cross-validation. Model interpretability was explored using SHapley Additive exPlanations to generate global and local explanations. An interview with a clinical expert assessed the utility of the explanations as the primary outcome, with trust in and expectations of both the model and the explanations as secondary outcomes.

**Results:** The model achieved an accuracy of 74.5%. Global explanations identified hip and knee weakness, gastrocnemius-soleus contractures, and BMI as the most important predictors across gait patterns. Local explanations illustrated how patient-specific impairments informed individual predictions. The user study demonstrated the clinical utility of the explanations, as they were perceived as interpretable, provided useful insights, and these insights were actionable. The explanations largely aligned with the expectations and increased self-reported trust in the model.

**Conclusions:** Gait patterns in DMD can be predicted from clinically actionable impairments, and explainable artificial intelligence can translate model outputs into meaningful clinical insights. This approach is promising for supporting both general and personalized rehabilitation and orthopedic strategies aimed at prolonging ambulation in DMD. Further validation in larger, multi-center cohorts is needed.

## 1. Background

‘Actionable’ impairments that contribute to gait pathology are targeted by rehabilitation and orthopedic strategies to prolong ambulation in children with Duchenne muscular dystrophy (DMD). DMD results from dystrophin deficiency, leading to progressive muscle degeneration and consequent impairments such as muscle weakness and contractures [1,2]. To data, there is still no cure. Research has mainly focused on pharmaceutical disease-modifying treatments, but their clinical implementation has been challenging [3–5]. Only corticosteroids have widely been accepted to slow disease progression [3,6,7], despite side effects such as weight gain [8,9], which also influences gait. Meanwhile, treatments addressing the underlying impairments remain understudied in DMD, despite their potential to impact gait decline. These impairments are ‘actionable’, meaning they are modifiable through targeted treatments. Advancing rehabilitation and orthopedic strategies therefore requires a comprehensive understanding of how actionable impairments contribute to gait pathology. However, this understanding remains limited.

Gait classifications can aid in clinical decision-making and improve communication among healthcare providers. Interpreting 3D gait analysis in DMD remains challenging due to the high dimensionality, temporal dependence, age-related variation, and heterogeneity in disease progression [10]. We therefore developed a DMD gait classification with a mildly affected gait pattern with only minor deviations, a tiptoeing gait pattern with the largest deviations at the ankle, and a flexion gait pattern with pronounced deviations at the trunk, pelvis and hip [11]. However, variability remains within these gait patterns, with some patients located at cluster boundaries and showing characteristics of multiple patterns, highlighting the need for personalized treatment strategies. To make gait classifications clinically useful for both general guidelines and personalized treatment strategies, it is essential to understand how actionable impairments contribute to gait patterns at both the general and patient-specific levels.

Unravelling the complex relationships between impairments and gait pathology requires advances computational approaches. Our previous work identified several relationships [12]. However, one-by-one associations cannot unravel the complex relationships and fails to reveal the most important contributors to the gait patterns. Predictive simulations based on musculoskeletal models and without relying on experimental data have shown that muscle weakness primarily caused gait deviations in DMD, while contractures further contributed to them, and that loss of ambulation was predicted when both impairments worsen together [13]. However, weakness and contractures of all muscle groups were simulated simultaneously, making it difficult to determine which muscle group contributed most. In addition, musculoskeletal models remain simplifications of the complex reality and cannot fully capture all experimentally observed deviations. Moreover, simulations were only evaluated at the group level using average models with ±1SD variations. Since DMD is highly heterogeneous, insights into individual predictions are important to support personalized treatment strategies. Therefore, to fully understand how actionable impairments contribute to the gait patterns at both the average and individual levels, a complementary data-driven modelling approach using machine learning is needed.

Machine learning techniques in 3D gait analysis [14,15] have been used for gait event detection [16], discrimination between typical and pathological gait [17], automated gait classification [18], treatment outcome predictions [19,20], and clinical decision support [21]. In DMD, however, their use remains limited [22–28]. To our knowledge, no study in DMD has used machine learning to predict gait patterns from actionable impairments.

Machine learning models have clinical value only if the predictions are understandable to clinicians. Explainable artificial intelligence (XAI) addresses this by revealing how input variables contribute to predictions, enabling actionable insights provided that these variables are ‘actionable’ themselves. XAI can generate both global and local explanations. Global explanations focus on the average contribution of features and can therefore facilitate the development of general treatment guidelines. In contrast, local explanations provide insights into individual predictions and enable personalized clinical decision-making and treatment strategies. Despite growing interest, XAI has seen limited application in gait analysis [14], and explanations linking actionable impairments to gait patterns are notably absent [15]. XAI is therefore required not only for transparency, but also to support clinically actionable decision-making for both general and personalized treatment strategies.

User studies with clinical experts are essential to identify actionable variables connecting to underlying impairments and to evaluate whether explanations provide clinical utility and increase trust in the model predictions. Yet, reviews report low methodological quality of existing studies regarding the appropriateness for clinical applications [15] and the need for more user-centered validation [29]. In cerebral palsy, two expert-validation studies [30,31] have demonstrated increased understanding, trust, and perceived usefulness of XAI. This highlights the importance of validation with clinical experts for translating insights from machine learning models into clinical decision-making.

The research questions of this study were: (1) Can gait patterns be accurately classified from actionable impairments? and (2) Can the model’s predictions be explained, and do these explanations provide clinical utility and increase trust in the model?

## 2. Methods

### 2.1 Retrospective dataset

The retrospective dataset used for this study is from the database of the Clinical Motion Analysis Laboratory of the University Hospital Leuven (CMAL-Leuven). The local ethics committee (Ethical Committee UZ Leuven/KU Leuven) approved this retrospective study (S71696). The data was collected between 2015 and 2022, and consists of instrumented strength assessment, clinical examination and 3D gait analysis. The dataset comprises 137 repeated measurements (multiple time points: one to ten, time interval between measurements: 5-35 months, total follow-up period: 6 months to 6 years) from 30 boys with DMD, resulting in 274 data points (two affected legs per measurement).

All data points were assigned to one of three gait patterns based on our DMD gait classification [11]. Domain experts clinically defined the patterns based on the cluster results and subsequently reviewed and reclassified the data points: mildly affected gait pattern (n = 171 legs, 85 measurements with both legs classified as mild and one mixed mild/tiptoeing measurement), tiptoeing gait pattern (n = 26 legs, 12 measurements with both legs classified as tiptoeing and two mixed measurements), and flexion gait pattern (n = 77 legs, 38 measurements with both legs classified as flexion gait and one mixed flexion/tiptoeing measurement). These final classifications served as the ground truth for training and evaluating the machine learning model.

The prediction set comprised 24 actionable impairments, defined as clinical features that can be directly modified by treatment (Table 1). These included muscle weakness, range of motion, muscle stiffness, and one subject characteristic.

**Table 1:** Overview of all features.

| Features | Type | Unit | Missing values (n) |
| --- | --- | --- | --- |
| **Muscle weakness** |  |  |  |
| Hip extension strength | numerical | % | 50 |
| Hip flexion strength | numerical | % | 50 |
| Hip abduction strength | numerical | % | 52 |
| Knee extension strength | numerical | % | 22 |
| Knee flexion strength | numerical | % | 22 |
| Dorsiflexion strength | numerical | % | 22 |
| Plantar flexion strength | numerical | % | 22 |
| Abdominal strength | ordinal | category | 18 |
| Back muscle strength | ordinal | category | 28 |
| **Range of motion** |  |  |  |
| Hip extension ROM | numerical | ° | 0 |
| Hip adduction ROM | numerical | ° | 87 |
| Knee extension ROM | numerical | ° | 0 |
| Hamstrings ROM | numerical | ° | 2 |
| Rectus femoris length | ordinal | category | 23 |
| Gastrocnemius ROM | numerical | ° | 0 |
| Soleus ROM | numerical | ° | 0 |
| **Muscle stiffness** |  |  |  |
| Hip flexor stiffness | ordinal | category | 158 |
| Hip abductor stiffness | ordinal | category | 158 |
| Knee flexor stiffness | ordinal | category | 158 |
| Hamstrings stiffness | ordinal | category | 160 |
| Rectus femoris stiffness | ordinal | category | 164 |
| Gastrocnemius stiffness | ordinal | category | 158 |
| Soleus stiffness | ordinal | category | 158 |
| **Subject characteristic** |  |  |  |
| Body mass index | numerical | kg/m<sup>2</sup> | 0 |

n: number; ROM: range of motion.

Muscle weakness was presented by nine features. Seven numerical features were obtained using fixed dynamometry [32,33]: hip extension, hip flexion, hip abduction, knee extension, knee flexion, dorsiflexion and plantar flexion. Strength was expressed as joint moments (Nm) and normalized to aged-matched typically developing reference values (%) [13,34]. Measurements were performed unilaterally. The assessed side was randomly selected by coin toss unless asymmetry (uncommon) was identified during manual muscle testing, in which case the weaker side was measured. Values were assigned bilaterally, because muscle weakness in DMD typically manifests symmetrically [35], and bilateral manual muscle testing showed little to no asymmetry in the present cohort. Two additional ordinal features, abdominal and back strength, were assessed using manual muscle testing [36]. As these measurements yield a single global score, the value was assigned bilaterally.

Seven range of motion (ROM) features were included. Six numerical features (degrees) were assessed using goniometry [37,38]: passive hip extension (modified Thomas test [38]), hip adduction (with hip and knee in 0° extension, and contralateral hip and knee in 90° flexion), knee extension, hamstrings ROM (true popliteal angle), gastrocnemius ROM (ankle dorsiflexion with knee extended), and soleus ROM (ankle dorsiflexion with knee flexed to 90°). In addition, one ordinal feature was included to categorize rectus femoris ROM (Duncan Ely test [39]: 0 = buttocks do not lift; 1 = buttocks lift at >100° knee flexion; 2 = buttocks lift between 80° and 100° knee flexion; and 3 = buttocks lift at <80° knee flexion). All ROM measures were obtained bilaterally.

Seven ordinal muscle stiffness features were assessed bilaterally during the passive ROM measurements using a clinical stiffness scale [13] (0=no increased resistance; 1= minimal increased resistance at the end of ROM; 2= increased resistance; 3= highly pronounced resistance): hip flexors, hip abductors, rectus femoris, knee flexors, hamstrings, gastrocnemius and soleus.

One numerical subject characteristic, i.e. body mass index (BMI), was included.

### 2.2 Preprocessing

Missing values in the muscle impairment variables were assumed to be missing at random and imputed using linear regression with age, BMI, and observed muscle impairments as predictors.

Multicollinearity was assessed using variance inflation factors and correlation matrices. Due to substantial multicollinearity, principal component analysis (PCA) was applied. To preserve interpretability, highly correlated muscle impairments were grouped into clinically meaningful categories, resulting in nine muscle impairment groups, while BMI was retained as a separate predictor. After standardization, PCA was performed within each group, and principal components (PCs) with eigenvalue >1 were retained (Table 2). This resulted in one PC per muscle impairment group. In total, 10 predictors were included: nine muscle impairment PCs and BMI.

**Table 2:** Overview of the principal components.

| Principal components | Explained variance | Muscle impairments | Loadings |
| --- | --- | --- | --- |
| Hip and knee weakness PC | 76.40% | Hip extension strength | -0.789 |
|  |  | Hip flexion strength | -0.878 |
|  |  | Hip abduction strength | -0.89 |
|  |  | Knee extension strength | -0.919 |
|  |  | Knee flexion strength | -0.799 |
| Ankle weakness PC | 69.10% | Dorsiflexion strength | -0.846 |
|  |  | Plantar flexion strength | -0.846 |
| Upper body weakness PC | 70.10% | Abdominal strength | -0.822 |
|  |  | Back muscle strength | -0.822 |
| Hip flexion contractures PC | 83.30% | Hip extension ROM | -0.916 |
|  |  | Hip flexor stiffness | 0.916 |
| Hip abduction contractures PC | 72.40% | Hip adduction ROM | -0.833 |
|  |  | Hip abductor stiffness | 0.833 |
| Knee flexion contractures PC | 91.70% | Knee extension ROM | -0.964 |
|  |  | Knee flexor stiffness | 0.964 |
| Hamstrings contractures PC | 78.60% | Hamstrings ROM | -0.869 |
|  |  | Hamstrings stiffness | 0.869 |
| Rectus femoris contractures PC | 66.30% | Rectus femoris length | 0.801 |
|  |  | Rectus femoris stiffness | 0.801 |
| Gastrocnemius and soleus contractures PC | 88.90% | Gastrocnemius ROM | -0.974 |
|  |  | Gastrocnemius stiffness | 0.913 |
|  |  | Soleus ROM | -0.929 |
|  |  | Soleus stiffness | 0.957 |

PC: principal component; ROM, range of motion.

All original features and PCs were scaled to a 0–1 range using Min-Max scaling.

### 2.3 Classification model

A random forest classifier was selected due to the small dataset, minimal hyperparameter tuning requirements, and good performance in related work [31]. Given the imbalanced class distribution, a balanced random forest classifier with class-balanced bootstrap sampling was used. Model performance was assessed using leave-one-group-out cross-validation with patient identity defining the groups, such that all datapoints from one patient were excluded from the training set and used as the test set in each fold, as in our previous study [11]. For evaluation, predicted class labels were assigned based on the highest class probability. Performance was summarized using confusion matrices, class-wise precision, recall, and F1-score, their weighted averages across gait patterns, and the overall accuracy.

### 2.4 Explanations

Since the random forest classifier is a black-box model, post-hoc XAI was applied using Tree SHapley Additive exPlanations (SHAP). For an individual prediction, SHAP values quantify both the magnitude and direction of each feature’s contribution, where the absolute SHAP value reflects the local feature importance and the sign indicates whether the feature increases (positive contribution) or decreases (negative contribution) the predicted probability. Both global and local explanations were generated. Before the global explanations were obtained, the model was retrained on the full dataset. Global explanations were then computed as mean absolute SHAP values, resulting in global feature importance scores. The entire study sample was used as the reference group for the global explanations. To illustrate the local explanations, two gait observations from the same patient were selected: observation A, classified as mildly affected, and observation B, classified as the flexion gait pattern. The model was retrained with both observations excluded, after which the SHAP values were computed for each observation. For the local explanations, the mildly affected gait pattern was used as the reference group, as maintaining this gait pattern is clinically the most relevant.

### 2.5 User study

A preliminary user study was conducted to validate the generated explanations. Following the recommendations of Davis et al.[40], utility was considered the primary evaluation outcome and trust a secondary outcome. Utility was assessed in terms of interpretability, usefulness and actionability. Trust in the model predictions and the insights of the explanations was measured using self-reported trust ratings on a 0–10 scale (0 = no trust; 10 = complete trust) before and after exposure to the explanations. As users’ prior expectations may influence trust [41], these expectations were also assessed.

To evaluate these outcomes, an online interview was conducted with one clinical expert who was familiar with the dataset but had no prior knowledge of the SHAP explanations used in this study. Before the interview, the expert received an overview of the data preprocessing, the development of the classification model, and its performance. Explanations of the SHAP method and its visualizations were introduced incrementally throughout the interview, allowing each visualization to be discussed immediately after its explanation. The interview followed a Think-Aloud protocol [42], in which the clinical expert verbalized their reasoning while completing a series of tasks. First, the PCs were evaluated with respect to their clinical interpretability and actionability. Next, observations A and B, previously introduced as the examples used to illustrate the local explanations, were presented to the expert, and the expert was asked to identify their gait patterns based on the PC values. Baseline trust in the model predictions on unseen data was then recorded before introducing the SHAP methodology. After explaining the SHAP methodology, the expert was asked whether the explanation of the methods and the selected reference population were clear and meaningful. The expert then completed two expectation-based tasks related to the global feature effects: (i) predicting the direction of the relationship (positive, negative, or no effect) between each actionable PC and the predicted probability of each gait pattern, and (ii) identifying the three PCs expected to have the greatest influence on the predictions. Subsequently, the predicted probabilities for observations A and B were presented, and the expert was asked whether these probabilities were considered plausible. Thereafter, three SHAP visualizations were presented sequentially. For each visualization, an explanation of its interpretation was first provided, followed by the same set of evaluation questions. Specifically, the expert was asked (i) which insights could be derived from the visualization, (ii) whether these insights were clinically actionable and how they could support clinical practice, (iii) whether the insights agreed with their prior expectations, and (iv) how easy the visualization was to interpret and use for obtaining insights. Finally, trust in the model predictions on unseen data and trust in the insights of the explanations were reassessed using the same 0–10 rating scales to evaluate changes following exposure to the explanations.

## 3. Results

### 3.1 Classification model: Random forest results

The results of the leave-one-group-out cross-validation are presented in Table 3. The accuracy of the model was 74.5%, as 204 of the 274 observations were correctly classified.

**Table 3.** Dataset composition by gait pattern, and patients per gait pattern (A), and leave-one-group-out cross-validation results: confusion matrix (B) and performance metrics (C)

| A |  | Mild<br>(n) | Tiptoeing<br>(n) | Flexion<br>(n) |
| --- | --- | --- | --- | --- |
| Legs (n) |  | 181 | 59 | 190 |
| Observations (n) |  | 91 | 17 | 45 |
| Patients (n) |  | 24 | 6 | 13 |

| B |  | Predicted |  |  |
| --- | --- | --- | --- | --- |
|  |  | Mild<br>(n) | Tiptoeing<br>(n) | Flexion<br>(n) |
| Actual | Mild (n) | 135 | 16 | 20 |
|  | Tiptoeing (n) | 7 | 16 | 3 |
|  | Flexion (n) | 15 | 9 | 53 |

| C |  | Precision<br>(%) | Recall<br>(%) | F<sub>1</sub> Score<br>(%) |
| --- | --- | --- | --- | --- |
| Mild |  | 86.0% | 78.9% | 82.3% |
| Tiptoeing |  | 39.0% | 61.5% | 47.8% |
| Flexion |  | 69.7% | 68.8% | 69.3% |
| Micro average |  | 74.5% | 74.5% | **74.5%** |
| Weighted average |  | 74.5% | 73.5% | **75.4%** |

n: number;

### 3.2 Explainability results

#### 3.2.1 Global explanations

After retraining on the full dataset, the model achieved an accuracy of 93.9%. Using the entire sample as reference, baseline predicted probabilities were 46.8%, 20.7% and 32.5% for the mildly affected, tiptoeing, and flexion patterns, respectively.

Global feature importance (mean absolute SHAP values) showed that the gastrocnemius and soleus contracture PC, the hip and knee weakness PC, and BMI were the most important predictors across gait patterns (Figure 1). The gastrocnemius and soleus contracture PC was most important for the mildly affected and tiptoeing gait patterns, and the hip and knee weakness PC for the flexion gait pattern.

**Figure 1.**
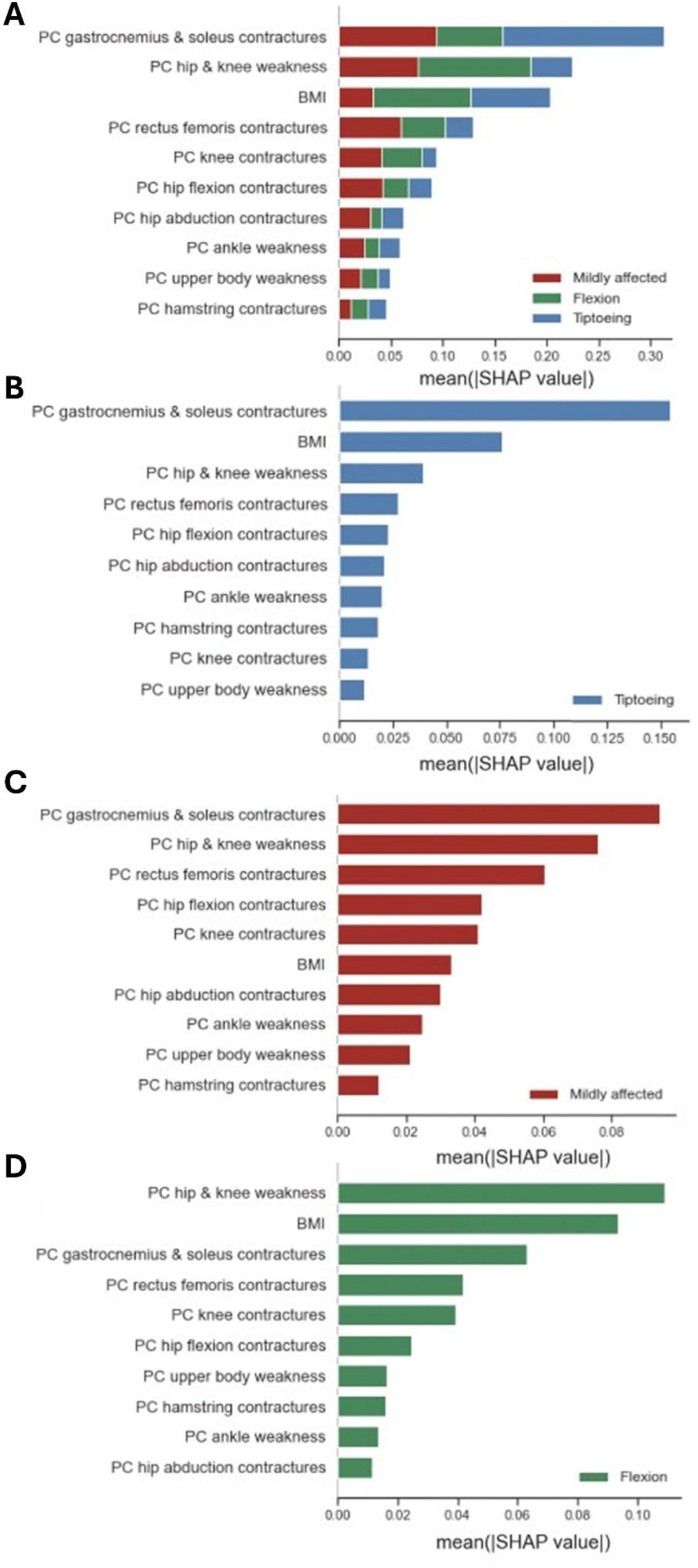
A. SHAP global feature importance over all gait patterns (**A**) and per gait pattern (**B,C,D**). Global feature importance is obtained from the mean absolute SHAP value per feature over all observations. The larger this value, the more important the feature is in the prediction. The features are sorted from most (top) to least (bottom) important.

Looking at the direction of feature contributions to gait pattern probabilities, low values across all features except the hamstrings contracture PC predicted the mildly affected gait pattern (Figure 2). High gastrocnemius and soleus contracture PC values, and low BMI and hip and knee weakness PC values contributed most to the tiptoeing gait pattern, whereas high hip and knee weakness PC values and BMI, and low gastrocnemius and soleus contracture PC values contributed most to the flexion gait pattern.

**Figure 2.**
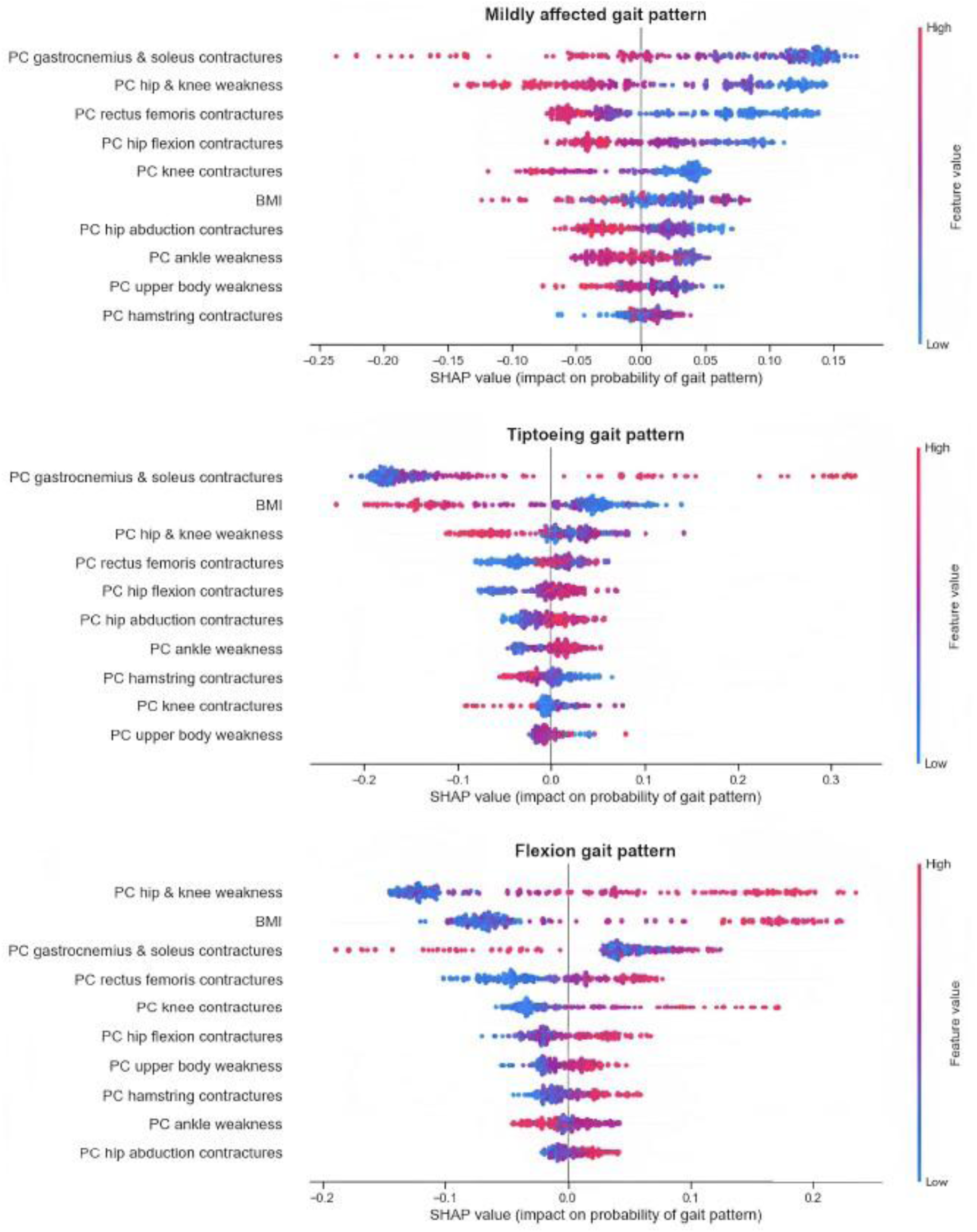
SHAP summary plots for each gait pattern. Features are ranked by importance. Each dot represents an observation, with color indicating the feature value (blue = high, pink = low). The position on the x-axis reflects the SHAP value associated with that feature value. Dots located farther from zero on the x-axis indicate a greater absolute effect of that feature on the prediction, which corresponds to the local feature importance.

#### 3.2.2 Local explanations

Local explanations are illustrated in this section using the two selected gait observations from the same patient as described above: gait observation A, classified as mildly affected, and gait observation B, classified as flexion.

The baseline prediction represents the model’s expected class probabilities before considering the features of the gait observations A and B. Using the mildly affected gait pattern as the reference group, these baseline class probabilities were 70.1%, 12.4%, and 17.5% for the mildly affected, tiptoeing, and flexion patterns, respectively. For both gait observation A and B, force plots were used to visualize how individual features shifted the model prediction from the baseline probability to the final class probability. For each gait pattern, features pushing the prediction above baseline indicate support for that pattern, whereas features pushing it below baseline indicate evidence against it. Only the most important feature contributions are described below.

For gait observation A, the predicted probabilities were 62%, 13%, and 25% for the mildly affected, tiptoeing, and flexion gait patterns, respectively. Thus, the observation was still classified as mildly affected, but compared with the baseline prediction, the model assigned less probability to mildly affected gait and more probability to the more advanced patterns. For the mildly affected pattern, higher rectus femoris contracture and hip–knee weakness PC values decreased probability, whereas lower hip flexion contracture PC and BMI values increased probability (Figure 3.A). For the tiptoeing pattern, lower gastrocnemius–soleus contracture PC values decreased probability, whereas lower trunk weakness PC values and higher rectus femoris contracture and hip–knee weakness PC values increased the probability (Figure 3.B). For the flexion pattern, higher rectus femoris contracture and hip–knee weakness PC values increased probability, whereas lower BMI decreased probability (Figure 3.C).

**Figure 3.**
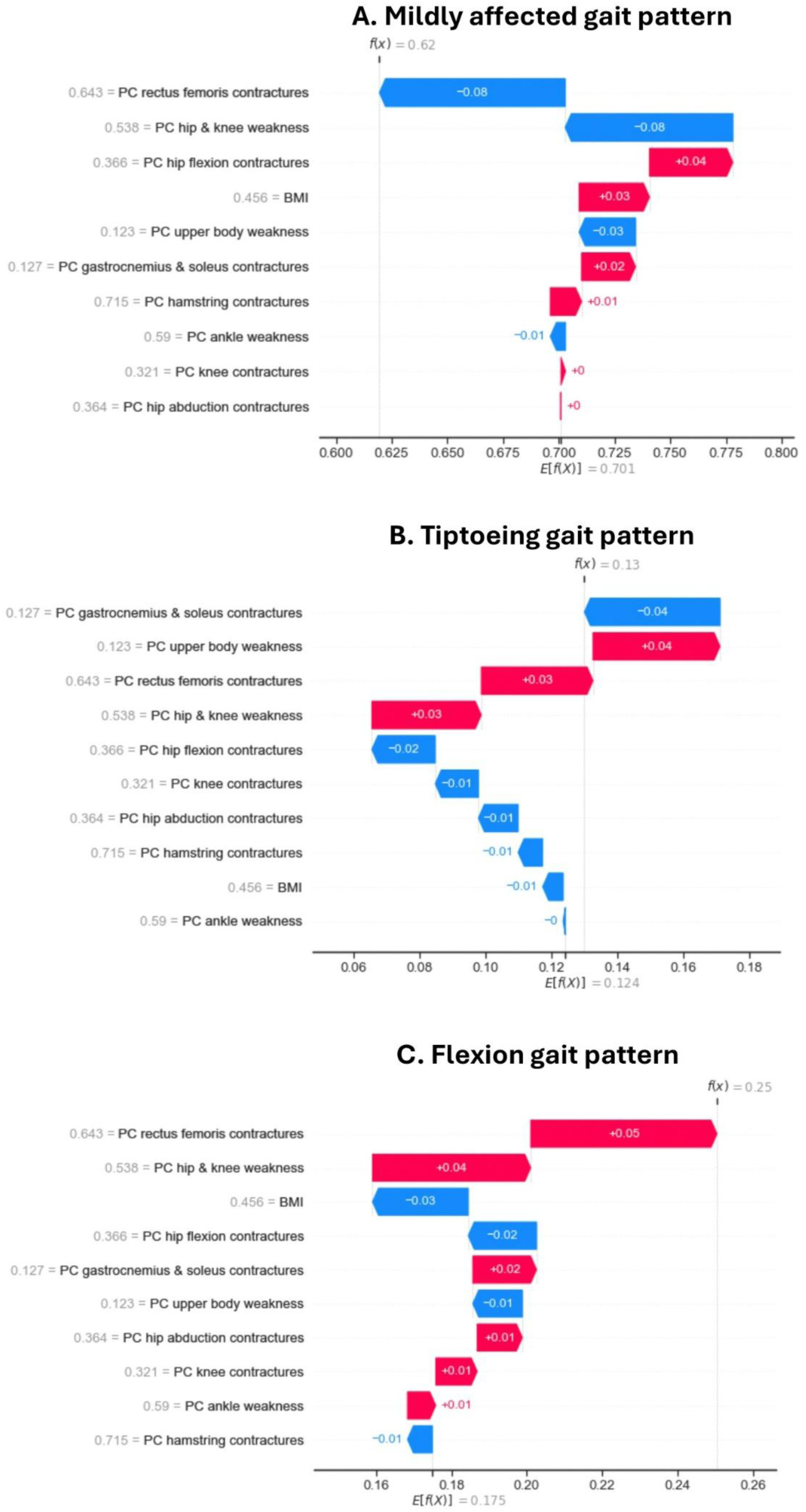
SHAP force plots for gait observation. **A**. The x-axis shows the baseline probability, E[f(X)], from which feature-specific SHAP values shift the prediction toward the final model output, f(X). Positive SHAP values (red) increase the predicted probability, whereas negative SHAP values (blue) decrease it. Features are ranked on the y-axis by local importance, quantified by the absolute SHAP value; arrow length represents effect magnitude and color indicates effect direction. The patient- and observation-specific feature value is shown next to each feature on a normalized scale from 0 (low value) to 1 (high value), allowing interpretation of whether a relatively low or high feature value contributes positively or negatively, and to what extent, to the predicted probability of the gait pattern.

For gait observation B, the predicted probabilities were 5%, 26% and 69% for the mildly affected, tiptoeing, and flexion gait patterns, respectively. Thus, the observation was classified as flexion gait pattern. Compared to the baseline prediction, the probability of the mildly affected gait pattern was much lower, and higher for the more advanced gait patterns. For the mildly affected pattern, higher gastrocnemius–soleus contracture PC, knee flexion contracture PC, and hip–knee weakness PC values decreased probability (Figure 4.A). For the flexion pattern, higher hip–knee weakness PC, knee flexion contracture PC, and BMI values increased probability, whereas higher gastrocnemius–soleus contracture PC values decreased probability (Figure 4.B).

**Figure 4.**
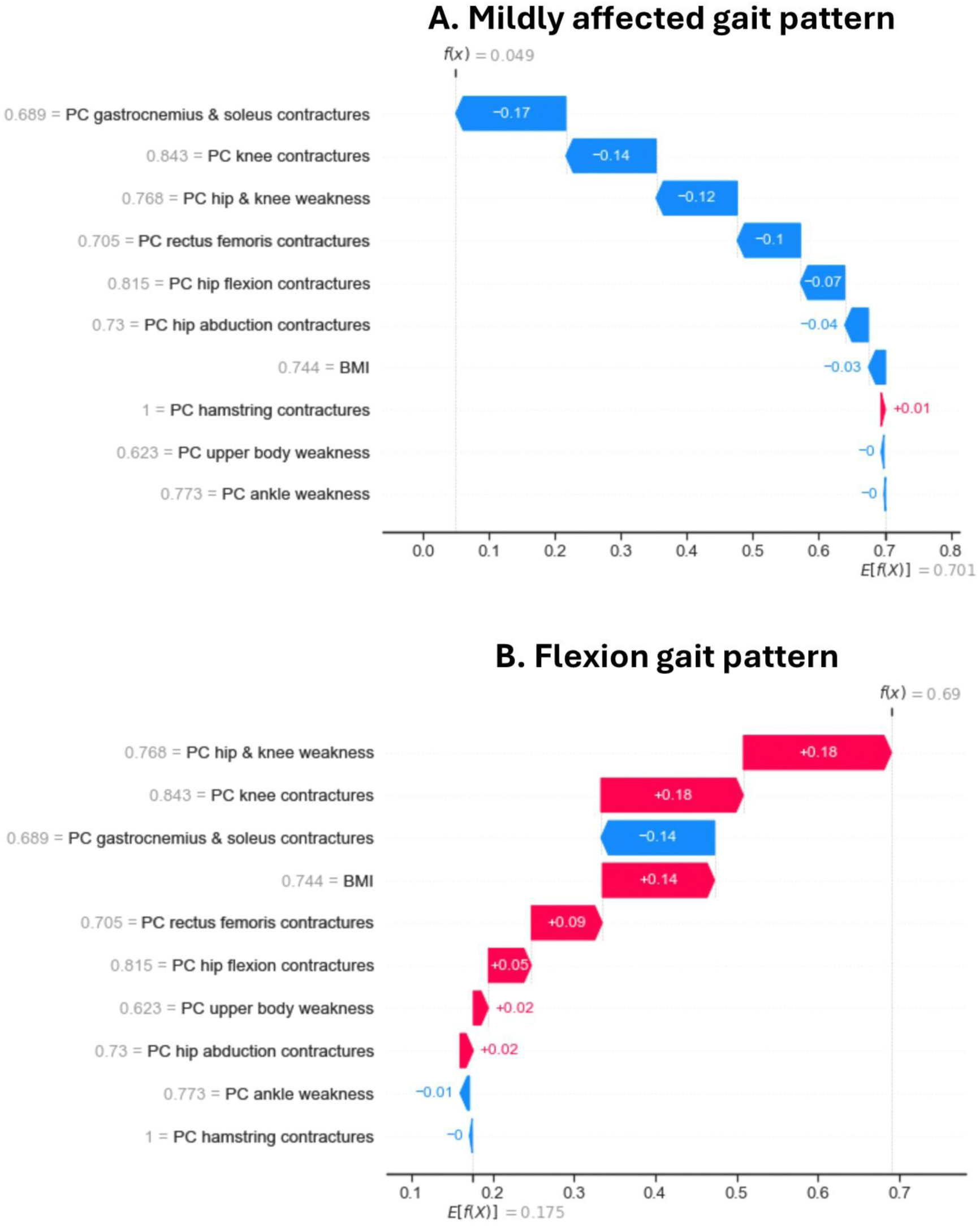
SHAP force plots for gait observation. **B**. The x-axis shows the baseline probability, E[f(X)], from which feature-specific SHAP values shift the prediction toward the final model output, f(X). Positive SHAP values (red) increase the predicted probability, whereas negative SHAP values (blue) decrease it. Features are ranked on the y-axis by local importance, quantified by the absolute SHAP value; arrow length represents effect magnitude and color indicates effect direction. The patient- and observation-specific feature value is shown next to each feature on a normalized scale from 0 (low value) to 1 (high value), allowing interpretation of whether a relatively low or high feature value contributes positively or negatively, and to what extent, to the predicted probability of the gait pattern.

### 3.3 User study: case study

The clinical expert indicated that the PCs had a clear clinical meaning and targeted intervention was possible for each feature. One suggestion was to distinguish between hip and knee weakness, as this distinction is clinically relevant: *“In the early phase, patients first present proximal weakness at the hip and only later at the knee.”*

The gait patterns of observations A and B were correctly identified based on the PC values by the clinical expert. Observation A was classified as mildly affected because of generally low feature values, although the relatively high value for the hamstring contracture PC introduced some uncertainty. Observation B was confidently classified as the flexion gait pattern because of its high feature values.

The SHAP methodology and the selected reference group were considered intuitive and well explained. The predicted probabilities for both observations were consistent with the expert’s expectations. The expert expected a relatively high probability for observation A to belong to the flexion gait pattern because of its high value on the hamstring contracture PC, and a higher probability for observation B to belong to the tiptoeing gait pattern than for observation A because of its high value on the gastrocnemius and soleus contracture PC.

Overall, the global explanations aligned well with the expert’s prior expectations. The most influential features identified by the global feature importance plot largely corresponded to the expected top predictors for each gait pattern, with only a few discrepancies (e.g., rectus femoris contracture PC was more important than expected by the clinical expert for the mildly affected gait pattern, whereas BMI, ankle weakness, and knee contractures were less important than anticipated for the mildly affected, tiptoeing, and flexion gait patterns, respectively). The participant considered the plot to be easily understandable, although the absence of information on the direction of feature effects made the plot harder to interpret.

Similarly, the summary plot largely confirmed the expected direction of the feature effects. Only two relationships appeared counter-intuitive: the positive relationship between the hamstrings contracture PC and the predicted probability for the mildly affected gait pattern and the negative relationship between the gastrocnemius and soleus contracture PC and the predicted probability for the flexion gait pattern. After discussing the latter in the context of competing gait pattern probabilities (i.e., tiptoeing gait pattern), the participant considered the explanation plausible. Although the summary plot required more effort to interpret than the feature importance plot, the participant indicated that this would improve with experience. Furthermore, the insights from both global visualizations could be translated into general actions in clinical practice which contributes to a higher level of utility.

The local force plots also showed a high degree of agreement with the expectations of the clinical expert, despite a few unexpected effects, most notably the limited and opposite effect of hamstring contracture PC for observation A. The clinical expert was also able to translate the local explanations into personalized clinical interventions for both observations. The clinical expert appreciated that the direction and magnitude of the feature contributions were clearly visualized, although interpreting the predictions relative to the reference group’s expected probability initially caused some confusion. Furthermore, because the displayed effects are tied to the observed feature values, the visualization does not directly indicate how changes in these feature values would affect the predicted probability. The clinical expert also noted that consulting the box plots was necessary to understand the position of a feature value within its distribution. Despite being more difficult to interpret than the global visualizations, the local explanations were considered more valuable because of their ability to support personalized treatment decisions: “*These local explanations have interesting implications as they help to personalize treatments. This is clinically highly relevant as DMD is a heterogeneous disease. It is also useful that this explainer takes the underlying interactions between the feature values into account because this is hard to take into consideration in clinical practice. For example, depending on BMI, two patients with the same muscle weakness may have a different effect of muscle weakness on the gait pattern which might result in different treatment decisions.*”

Self-reported trust in the model increased throughout the interview, from the initial assessment based solely on the model performance, to after reviewing the predicted probabilities, and finally after evaluating the global and local explanations. According to the clinical expert, this increase in trust resulted from the strong correspondence between the explanations and their clinical expectations, with trust expected to increase further as the model demonstrates its reliability in clinical practice.

## 4. Discussion

In this study, DMD gait patterns were classified from clinically actionable impairments with good accuracy. Global explanation identified hip and knee weakness, gastrocnemius and soleus contractures, and BMI as the most important predictors, informing general clinical guidelines. A case-based example illustrated how local explanations can reveal patient-specific drivers of the predicted gait pattern, highlighting the potential for personalized clinical decision-making. Finally, a preliminary user study with a clinical expert indicated the clinical utility of the explanations and increased the expert’s self-reported trust in the model.

The actionable impairments were able to classify DMD gait patterns with an accuracy of 75%. A random forest model was selected due to its demonstrated robustness in previous work [31] and its favorable performance in smaller datasets, with a reduced risk of overfitting. Although overall performance was good, classification of the tiptoeing gait pattern during cross-validation was poor, most likely due to the very small and imbalanced sample size, which was dominated by a single participant. The achieved overall accuracy is lower than that reported in several machine learning studies predicting gait parameters or gait patterns, where accuracies exceeding 90% are frequently reported [17,31,43–48], including the 91% accuracy for the DMD gait classification by Sutherland et al.[25]. However, these higher accuracies are typically achieved by using 3D gait data, or a combination of 3D gait data and direct measurements of the underlying impairments (e.g., ROM measured with goniometry, muscle weakness measured via fixed dynamometry), rather than impairment data alone. Studies relying on clinical impairment measures have shown poorer performance [49] than observed in the present study. Since gait pathology arises from underlying impairments that are the primary targets of rehabilitation and orthopedic interventions, the ability to predict DMD gait patterns based solely on clinically actionable impairments, with good accuracy, may support clinical decision-making by promoting transitions toward milder gait patterns or preventing progression to more severely affected gait patterns.

The explanations of the model provided clinically actionable insights into the underlying impairments predicting DMD gait patterns, informing population-level treatment strategies and general clinical guidelines. At the population level, global explanations identified gastrocnemius and soleus contractures, hip and knee weakness, and BMI as key predictors, highlighting these factors as central targets for rehabilitation and orthopedic management. As expected, overall lower impairment severity was important for the mildly affected gait pattern, indicating that reducing impairments helps maintaining the mildly affected gait pattern and preventing progression toward more affected gait patterns. For the tiptoeing gait pattern, increased gastrocnemius and soleus contractures emerged as the most influential predictors, emphasizing the importance of targeting ankle contractures through interventions such as stretching, night-time ankle–foot orthoses or serial casting for this gait pattern. For the flexion gait pattern, increased hip and knee weakness and increased BMI were the most important predictors, indicating that maintaining hip and knee strength through physiotherapy or additional passive support via orthoses during gait, and reducing BMI through dietary interventions are particularly relevant strategies for this gait pattern. Remarkably, BMI emerged as a highly influential factor, which was previously hypothesized in our earlier work [11] and now empirically supported, underscoring the important role of weight management in the prevention of progressing toward more affected gait patterns. Knee flexion contractures occurred less frequently, but when present were associated with an increased likelihood of a flexion gait pattern, whereas decreased hip flexion contractures were associated with an increased likelihood of the mildly affected gait pattern, and increased values with both the tiptoeing and flexion gait patterns. Together, these global explanations highlight specific actionable targets that can inform general rehabilitation and orthopedic guidelines tailored to distinct DMD gait patterns.

These findings are consistent with previously identified relationships [12], where hip extension, hip abduction and knee extension weakness was strongly associated with gait characteristics of the flexion pattern such as posterior trunk lean, anterior pelvic tilt, increased step width, and altered foot progression, while gastrocnemius and soleus contractures were linked to gait characteristics of the tiptoeing gait pattern such as reduced ankle dorsiflexion at initial contact and during swing. Although knee extension weakness has previously been associated with reduced dorsiflexion at initial contact [12], the present results indicate that ankle contractures play a more prominent role than weakness in predicting the tiptoeing gait pattern. This contrasts with our earlier simulation work [13], in which tiptoeing already emerged from simulating weakness alone. Notably, ankle weakness did not show a relationship with gait characteristics [12] and did not emerge as an important predictor in the current study.

Beyond these population-level insights, local explanations demonstrated the potential for personalized clinical decision-making by identifying patient- and observation-specific drivers of gait pathology. For example, when the presented case exhibited the mildly affected gait pattern, rectus femoris tightness and hip and knee weakness predominantly increased the probability of a flexion gait pattern, indicating the need for targeted stretching and strengthening interventions. In a subsequent gait observation when the case exhibited the flexion gait pattern, increased hip and knee weakness, increased knee flexion contracture, and higher BMI were the main contributors. While maintaining hip and knee strength remained an important personalized treatment goal, the progressive nature of DMD limits the ability to prevent progressive muscle weakness, warranting consideration of orthotic solutions providing passive support at the hip and knee. Although knee flexion contractures were less influential at the population level for this gait pattern, they were clinically highly relevant for this individual and therefore required prioritization through targeted stretching, positioning, and potential surgical consultation, particularly given that significant contractures may limit the feasibility of orthotic interventions. In addition, the increased contribution of BMI underscores the importance of individualized dietary management [8]. These findings illustrate how explainable machine-learning models can support personalized treatment planning in DMD gait management.

The preliminary user study suggested that the proposed explanations have potential clinical utility. Both the global and local explanations were perceived as interpretable, provided useful insights, and these insights were actionable. Notably, global explanations were easier to interpret and supported general treatment planning, whereas local explanations were more challenging to understand, despite their particular value for personalized decision-making. This suggests that future work should focus on enhancing the interpretability and presentation of local explanations, for instance through interactive or visual dashboard tools [50,51], to improve their clinical utility. The global and local explanations were also largely consistent with the clinical expert’s expectations, providing an indication that the explanations reflected existing clinical reasoning. This alignment was accompanied by an increase in self-reported trust in the model following exposure to the explanations, suggesting that explanations consistent with users’ mental models may foster trust in AI-supported clinical decision-making. These findings address concerns raised in previous reviews, which highlighted the limited appropriateness for clinical applications [15] and emphasized the need for more user-centered validation [29]. They are also consistent with earlier studies in cerebral palsy [30,31], which reported improved trust and perceived utility following expert evaluation of XAI, supporting the relevance of expert validation. Finally, explanations that confirm existing clinical knowledge are valuable for increasing trust in model predictions, but their greatest potential lies in uncovering reliable, previously unknown patterns. The clinical relevance of such discoveries can only be established through long-term evaluation and application in clinical practice. Although further validation involving multiple clinical experts is required, these preliminary findings indicate that the proposed framework has potential clinical utility and may facilitate the acceptance and translation of the model into clinical practice.

This study has some limitations. The dataset was small and affected by class imbalance. Therefore, a balanced random forest was used, as this model performs well in small datasets and mitigates imbalance by drawing bootstrap samples with equal class distributions. However, the limited sample size prevented the use of a separate validation set for hyperparameter tuning. In particular, the tiptoeing gait pattern was represented by a very small sample (n = 26 legs) and was dominated by a single participant. Excluding this participant during cross-validation resulted in poor performance for this class. In addition, high correlations among predictors required dimensionality reduction using PCA, which limited the ability to disentangle the contribution of individual clinical impairments (e.g., specific muscle weaknesses). Future work should explore and compare different XAI approaches to further improve interpretability. Finally, the user study included only one clinical expert. Larger studies with multiple experts and quantitative analyses are needed to draw stronger conclusions.

## 5. Conclusion

This study showed that DMD gait patterns can be accurately predicted from actionable impairments and that XAI can link model outputs to clinical interpretation. By combining classification with global and patient-specific explanations, the proposed framework improved understanding of how underlying impairments contribute to gait pathology, supporting both general clinical guidelines and personalized treatment strategies. Importantly, the preliminary user study demonstrated the potential clinical utility of the explanations and increased a clinical expert’s trust in the model, thereby addressing a key barrier to clinical adoption. Together, these findings suggest that this explainable, impairment-based approach has clear potential to enhance clinical decision-making and advance rehabilitation and orthopedic management aimed at prolonging ambulation in DMD. Future work should focus on validation in larger, multi-center cohorts and on involving a broader group of clinicians to further assess clinical usability and generalizability.

## Data Availability

All data concerning this study is available within the manuscript. Detailed data is available upon reasonable request to the first author.

## List of abbreviations

BMI: body mass index
CMAL-Leuven: Clinical Motion Analysis Laboratory of the University Hospital Leuven campus Pellenberg
DMD: Duchenne muscular dystrophy
PCA: principal component analysis
PC: principal component
ROM: range of motion
SHAP: SHapley Additive exPlanations
XAI: Explainable artificial intelligence

## Declarations

### Ethics approval and consent to participate

Data collection was approved by the local ethics committee (Ethical Committee UZ Leuven/KU Leuven; S71696) under the Declaration of Helsinki. All methodology adhered to the relevant regulations and guidelines.

### Consent for publication

Not applicable.

### Authors’ contributions

The study was conceptualized and designed by IV and TDL. Data analysis and visualizations were carried out by IV, BL and TDL. The user study was carried out by IV and BL. IV, BL and TDL interpreted the results. The original draft was written by IV, while TDL reviewed and edited the manuscript. All authors have read and approved the published version of the manuscript.

## References

1. Sussman M. Duchenne Muscular Dystrophy. J Am Acad Orthop Surg. 2002;10:138–51. 10.5435/00124635-200203000-00009

2. Duan D, Goemans N, Takeda S, Mercuri E, Aartsma-Rus A. Duchenne muscular dystrophy. Nat Rev Dis Prim. 2021;7:13. 10.1038/s41572-021-00248-3

3. Markati T, Oskoui M, Farrar MA, Duong T, Goemans N, Servais L. Emerging therapies for Duchenne muscular dystrophy. Lancet Neurol [Internet]. Elsevier Ltd; 2022;21:814–29. 10.1016/S1474-4422(22)00125-9

4. Ricci G, Bello L, Torri F, Schirinzi E, Pegoraro E, Siciliano G. Therapeutic opportunities and clinical outcome measures in Duchenne muscular dystrophy. Neurol Sci [Internet]. Springer International Publishing; 2022;43:625–33. 10.1007/s10072-022-06085-w

5. Goemans N. Therapy development and clinical outcome measures for Duchenne muscular dystrophy [PhD thesis]. Leuven: KU Leuven; 2013.

6. Birnkrant DJ, Bushby K, Bann CM, Apkon SD, Blackwell A, Brumbaugh D, et al. Diagnosis and management of Duchenne muscular dystrophy, part 1: diagnosis, and neuromuscular, rehabilitation, endocrine, and gastrointestinal and nutritional management. Lancet Neurol. 2018. p. 251–67. 10.1016/S1474-4422(18)30024-3

7. Birnkrant DJ, Bushby K, Bann CM, Alman BA, Apkon SD, Blackwell A. Diagnosis and management of Duchenne muscular dystrophy, part 2: respiratory, cardiac, bone health, and orthopaedic management. Lancet Neurol. 2018;17:347–61. doi:10.1016/S1474-4422(18)30025-5

8. Weber DR, Hadjiyannakis S, McMillan HJ, Noritz G, Ward LM. Obesity and endocrine management of the patient with Duchenne muscular dystrophy. Pediatrics. 2018;142:S43–52. 10.1542/peds.2018-0333F

9. Lamb MM, West NA, Ouyang L, Yang M, Weitzenkamp D, James K, et al. Corticosteroid treatment and growth patterns in ambulatory males with duchenne muscular dystrophy. J Pediatr. 2016;173:207–13. 10.1016/j.jpeds.2016.02.067

10. Vandekerckhove I, Van den Hauwe M, De Beukelaer N, Stoop E, Goudriaan M, Delporte M, et al. Longitudinal Alterations in Gait Features in Growing Children With Duchenne Muscular Dystrophy. Front Hum Neurosci. 2022;16:861136. 10.3389/fnhum.2022.861136

11. Vandekerckhove I, Papageorgiou E, Hanssen B, De Beukelaer N, Van den Hauwe M, Goemans N, et al. Gait classification for growing children with Duchenne muscular dystrophy. Sci Rep. 2024;14:10828. doi: 10.1038/s41598-024-61231-y

12. Vandekerckhove I, Molenberghs G, Van den Hauwe M, Goemans N, De Waele L, Van Campenhout A, et al. Longitudinal interaction between muscle impairments and gait pathology in growing children with Duchenne muscular dystrophy. J Neuroeng Rehabil. 2025;22:207. 10.1186/s12984-025-01718-5

13. Vandekerckhove I, D’Hondt L, Gupta D, Van Den Bosch B, Van den Hauwe M, Goemans N, et al. Muscle weakness but also contractures contribute to the progressive gait pathology in children with Duchenne muscular dystrophy: a simulation study. J Neuroeng Rehabil [Internet]. BioMed Central; 2025;22:103. 10.1186/s12984-025-01631-x

14. Dibbern KN, Krzak MG, Olivas A, Albert M V., Krzak JJ, Kruger KM. Scoping Review of Machine Learning Techniques in Marker-Based Clinical Gait Analysis. Bioengineering. 2025;12:591. 10.3390/bioengineering12060591

15. Samadi Kohnehshahri F, Merlo A, Mazzoli D, Bò MC, Stagni R. Machine learning applied to gait analysis data in cerebral palsy and stroke: A systematic review. Gait Posture [Internet]. Elsevier B.V.; 2024;111:105–21. 10.1016/j.gaitpost.2024.04.007

16. Kim YK, Visscher RMS, Viehweger E, Singh NB, Taylor WR, Vogl F. A deep-learning approach for automatically detecting gait-events based on foot-marker kinematics in children with cerebral palsy-Which markers work best for which gait patterns? PLoS One [Internet]. 2022;17:e0275878. 10.1371/journal.pone.0275878

17. Slijepcevic D, Horst F, Lapuschkin S, Horsak B, Raberger A-M, Kranzl A. Explaining Machine Learning Models for Clinical Gait Analysis. ACM Trans Comput Healthc. 2021;3:14. 10.1145/3474121

18. Papageorgiou E, Nieuwenhuys A, Vandekerckhove I, Van Campenhout A, Ortibus E, Desloovere K. Systematic review on gait classifications in children with cerebral palsy: an update. Gait Posture [Internet]. Elsevier; 2019;69:209–23. 10.1016/j.gaitpost.2019.01.038

19. Schwartz MH, Rozumalski A, Truong W, Novacheck TF. Predicting the outcome of intramuscular psoas lengthening in children with cerebral palsy using preoperative gait data and the random forest algorithm. Gait Posture [Internet]. Elsevier B.V.; 2013;37:473–9. 10.1016/j.gaitpost.2012.08.016

20. Schwartz MH, Rozumalski A, Novacheck TF. Femoral derotational osteotomy: Surgical indications and outcomes in children with cerebral palsy. Gait Posture [Internet]. Elsevier B.V.; 2014;39:778–83. 10.1016/j.gaitpost.2013.10.016

21. Chia K, Fischer I, Thomason P, Graham HK, Sangeux M. A Decision Support System to Facilitate Identification of Musculoskeletal Impairments and Propose Recommendations Using Gait Analysis in Children With Cerebral Palsy. Front Bioeng Biotechnol. 2020;8:529415. 10.3389/fbioe.2020.529415

22. Ramli AA, Liu X, Berndt K, Goude E, Hou J, Kaethler LB, et al. Gait Characterization in Duchenne Muscular Dystrophy (DMD) Using a Single-Sensor Accelerometer: Classical Machine Learning and Deep Learning Approaches. Sensors. 2024;24:1123. 10.3390/s24041123

23. Ramli AA, Liu X, Berndt K, Chuah C, Goude E, Kaethler LB, et al. Gait Event Detection and Travel Distance Using Waist-Worn Accelerometers across a Range of Speeds : Automated Approach. Sensors [Internet]. 2024;24:1155. 10.3390/s24041155

24. Ali MM, Hassan MM, Zaki M. Human Pose Estimation for Clinical Analysis of Gait Pathologies. Bioinform Biol Insights [Internet]. 2024;18:1–17. 10.1177/11779322241231108

25. Sutherland DH, Olshen R, Cooper L, Wyatt M, Leach J, Mubarak S, et al. The pathomechanics of gait in Duchenne Muscular Dystrophy. Dev Med Child Neurol. 1981;23:3–22. 10.1111/j.1469-8749.1981.tb08442.x

26. Sienko Thomas S, Buckon CE, Nicorici A, Bagley A, McDonald CM, Sussman MD. Classification of the gait patterns of boys with Duchenne muscular dystrophy and their relationship to function. J Child Neurol. 2010;25:1103–9. 10.1177/0883073810371002

27. Ricotti V, Kadirvelu B, Selby V, Festenstein R, Mercuri E, Voit T, et al. Wearable full-body motion tracking of activities of daily living predicts disease trajectory in Duchenne muscular dystrophy. Nat Med. Springer US; 2023;29:95–103. 10.1038/s41591-022-02045-1

28. Henricson EK, Ramli AA. Harnessing Fast Fourier Transform for Rapid Community Travel Distance and Step Estimation in Children with Duchenne Muscular Dystrophy. Sensors. 2025;25:3234. 10.3390/s25103234

29. Xiang L, Gao Z, Yu P, Fernandez J, Gu Y, Wang R, et al. Explainable artificial intelligence for gait analysis: advances, pitfalls, and challenges - a systematic review. Front Bioeng Biotechnol. 2025;13:1671344. 10.3389/fbioe.2025.1671344

30. Rind A, Slijepčević D, Zeppelzauer M, Unglaube F, Kranzl A, Horsak B. Trustworthy Visual Analytics in Clinical Gait Analysis: A Case Study for Patients with Cerebral Palsy. Proc - 2022 IEEE Work Trust Expert Vis Anal TREX 2022. 2022; 10.1109/TREX57753.2022.00006

31. Slijepcevic D, Zeppelzauer M, Unglaube F, Kranzl A, Breiteneder C, Horsak B. Explainable Machine Learning in Human Gait Analysis: A Study on Children With Cerebral Palsy. IEEE Access. IEEE; 2023;11:65906–23. 10.1109/ACCESS.2023.3289986

32. Goudriaan M, Nieuwenhuys A, Schless S, Goemans N, Molenaers G, Desloovere K. A new strength assessment to evaluate the association between muscle weakness and gait pathology in children with cerebral palsy. PLoS One. 2018;13:e0191097. 10.1371/journal.pone.0191097

33. Verreydt I, Vandekerckhove I, Stoop E, Peeters N, van Tittelboom V, Van de Walle P, et al. Instrumented strength assessment in typically developing children and children with a neural or neuromuscular disorder: A reliability, validity and responsiveness study. Front Physiol. 2022;13:855222. 10.3389/fphys.2022.855222

34. Vandekerckhove I, Hanssen B, Peeters N, Dewit T, De Beukelaer N, Van den Hauwe M, et al. Anthropometric-related percentile curves for muscle size and strength of lower limb muscles of typically developing children. J Anat [Internet]. 2025;00:1–15. 10.1111/joa.14241

35. Darras BT, Urion DK, Ghosh PS. Dystrophinopathies 2000 Sep 5 [Updated 2022 Jan 20]. In: Adam MP, Feldman J, Mirzaa GM, et al., editors. GeneReviews® [Internet]. Seattle (WA): University of Washington, Seattle; 1993-2025.

36. Hislop H, Avers D, Brown M. Muscle Testing Techniques of Manual Examination. 9th ed. Philadelphia. Elsevier. 1995;

37. Vandekerckhove I, Van den Hauwe M, Dewit T, Molenberghs G, Goemans N, De Waele L, et al. Longitudinal trajectories of muscle impairments in growing boys with Duchenne muscular dystrophy. PLoS One [Internet]. 2025;20:e0307007. 10.1371/journal.pone.0307007

38. Mudge AJ, Bau K V., Purcell LN, Wu JC, Axt MW, Selber P, et al. Normative reference values for lower limb joint range, bone torsion, and alignment in children aged 4-16 years. J Pediatr Orthop Part B. 2014;23:15–25. 10.1097/BPB.0b013e328364220a

39. Olivencia O, Godinez GM, Dages J, Duda C, Kaplan K, Kolber MJ, et al. the Reliability and Minimal Detectable Change of the Ely and Active Knee Extension Tests. Int J Sports Phys Ther. 2020;15:776–82. 10.26603/ijspt20200776

40. Davis FD. Perceived usefulness, perceived ease of use, and user acceptance of information technology. MIS Q. 1989;13:319–39. 10.2307/249008

41. Ooge J, Verbert K. Trust in Prediction Models: a Mixed-Methods Pilot Study on the Impact of Domain Expertise. 2021 IEEE Work Trust Expert Vis Anal TREX. IEEE; 2021;8–13. 10.1109/TREX53765.2021.00007

42. Jääskeläinen R. Think-aloud protocol. Handbook of translation studies. 2010.

43. Lan Z, Lempereur M, Gueret G, Houx L, Cacioppo M, Pons C, et al. Towards a diagnostic tool for neurological gait disorders in childhood combining 3D gait kinematics and deep learning. Comput Biol Med [Internet]. Elsevier Ltd; 2024;171:108095. 10.1016/j.compbiomed.2024.108095

44. Eskofier BM, Federolf P, Kugler PF, Nigg BM. Marker-based classification of young – elderly gait pattern differences via direct PCA feature extraction and SVMs. Comput Methods Biomech Biomed Engin. 2013;4:435–442. 10.1080/10255842.2011.624515

45. Begg R, Kamruzzaman J. A Comparison of Neural NetFvorks and Support Vector Machines for Recognizing Young-Old Gait Patterns. Conf Converg Technol Asia-Pacific Reg TENCON 2003, [Internet]. 2003;1:354–358. 10.1109/TENCON.2003.1273344

46. Wu J, Wang J, Liu L. Feature extraction via KPCA for classification of gait patterns. Hum Mov Sci. 2007;26:393–411. 10.1016/j.humov.2007.01.015

47. Zhou Y, Romijnders R, Hansen C, Campen J Van, Maetzler W, Hortobágyi T, et al. The detection of age groups by dynamic gait outcomes using machine learning approaches. Sci Rep. 2020;10:4426. 10.1038/s41598-020-61423-2

48. Bajpai R, Tiwari A, Joshi D, Khatavkar R. AbnormNet : A Neural Network Based Suggestive Tool for Identifying Gait Abnormalities in Cerebral Palsy Children. 2022 Int Conf Adv Technol ICONAT [Internet]. IEEE; 2022;1–5. 10.1109/ICONAT53423.2022.9725832

49. Ploeger HE, Bus SA, Nollet F, Brehm M. Gait patterns in association with underlying impairments in polio survivors with calf muscle weakness. Gait Posture [Internet]. Elsevier; 2017;58:146–53. 10.1016/j.gaitpost.2017.07.107

50. Spinner T, Schlegel U, Schäfer H, El-Assady M. ExplAIner: A Visual Analytics Framework for Interactive and Explainable Machine Learning. IEEE Trans Vis Comput Graph. 2020;26:1064–74. 10.1109/TVCG.2019.2934629

51. Caban JJ, Gotz D. Visual analytics in healthcare - opportunities and research challenges. J Am Med Informatics Assoc. 2015;22:260–2. 10.1093/jamia/ocv006

